# Differential connectivity of frontolimbic circuit induced by individualized disorder-specific stimuli in distinct symptom profiles of obsessive-compulsive disorder

**DOI:** 10.1101/2024.02.16.24302954

**Authors:** Navya Spurthi Thatikonda, Janardhanan C Narayanaswamy, Ganesan Venkatasubramanian, Janardhan Reddy Y. C., Shyam Sundar Arumugham

## Abstract

**Background:** Emotional processing deficits in obsessive-compulsive disorder (OCD) are reportedly caused by an aberrant frontolimbic circuit activation with inconsistent evidence, possibly due to symptom heterogeneity. We compared the activation and connectivity patterns of the frontolimbic structures during symptom provocation between patients with distinct symptom profiles of OCD.

**Methods:** We recruited 37 symptomatic OCD subjects categorized based on predominant symptom profiles, i.e., 19 with contamination/washing symptoms (OCD-C) and 18 with taboo thoughts (OCD-T), along with 17 healthy controls (HC). All subjects were evaluated with comprehensive clinical assessments and functional magnetic resonance imaging (fMRI) while appraising personalized disorder-specific visual stimuli with contrasting neutral stimuli as part of an individualized symptom provocation (ISP) task.

**Results:** OCD-C subjects had decreased task-dependent mean activation of left amygdala and right hippocampus compared to the other groups during symptom induction. Task-modulated functional connectivity analyses during ISP task revealed that HC had increased connectivity between right hippocampus and bilateral supplementary motor cortex, right insula and left cerebellum, left insula and inferior temporal gyrus than OCD-C. OCD-T subjects had greater connectivity between right insula and left cerebellum than OCD-C and increased connectivity of medial frontal cortex with right lateral occipital cortex than HC.

**Conclusions:** Contamination-related symptoms had decreased activation and altered connectivity of frontolimbic circuit during emotional provocation. Replicating these findings on larger samples with other symptom profiles might help develop personalized, targeted interventions for this heterogeneous disorder.

**Highlights:** OCD subjects with contamination symptom dimension had decreased activity in the hippocampus and amygdala and abnormal connectivity of middle frontal, hippocampus, and insular cortex with areas beyond the established CSTC circuit during individualized symptom provocation. In contrast, symptom provocation in taboo thoughts subjects revelead enhanced connectivity of limbic structures with visual processing areas. These findings sustain the notion of unique neurobiological correlates of the symptom dimensions, which needs to be substantiated in larger and homogenous samples with inclusion of other symptom profiles to further improve our insight into this heterogeneous disorder.

## 1. Introduction

Despite its current nosological status as a unitary entity, obsessive-compulsive disorder (OCD) shows considerable heterogeneity in clinical profile, course of illness, and treatment response (Lochner and Stein 2003). Factor analytic studies have found temporally and cross-culturally stable sets of symptom dimensions, including symmetry/arranging, aggression, forbidden/taboo thoughts, contamination/cleaning, and hoarding (Baer 1994; Batistuzzo et al. 2021; Bloch et al. 2008; Mataix-Cols et al. 2002). These symptom profiles are correlated with distinctive clinical manifestations in terms of age at onset, comorbidities, degree of insight, and neuropsychological deficits, supporting their validity (Prabhu et al. 2013; H. Kashyap et al. 2017; Kichuk et al. 2013). Recent neuroimaging studies have shown that the standard cortico-striato-thalamo-cortical (CSTC) model alone may be inadequate to explain this heterogeneity (Milad and Rauch 2012) and indicated evidence for the involvement of wider cortical and subcortical structures beyond the CSTC circuit (Menzies et al. 2008; van den Heuvel et al. 2016). It has been postulated that the diverse clinical presentations and the associated neurocognitive deficits may be explained by the varied involvement of the dorsal cognitive, ventral cognitive, fronto-limbic, sensorimotor, and frontoparietal networks (Shephard et al. 2021; 2022).

Dysregulated fear, anxiety, or distress is often seen in OCD patients with an attentional bias toward stimuli with negative valence (Kuelz, Hohagen, and Voderholzer 2004; Melli et al. 2017; Rao et al. 2010; Schienle et al. 2005; Moritz et al. 2009). Frontolimbic circuit comprising of amygdala, ventromedial prefrontal cortex including ventral anterior cingulate cortex, and hippocampus is implicated in the generation and evaluation of emotional responses, with a top-down control exerted by dorsal prefrontal cortex to regulate the generated emotions (Shephard et al. 2021; 2022). Symptom provocation tasks designed using disorder-specific stimuli (with varying grades of idiosyncrasy) and contrasting non-specific (general aversive or neutral) stimuli are commonly employed in functional magnetic resonance imaging (fMRI) studies to study emotional processing deficits (De Putter, Van Yper, and Koster 2017). OCD patients have shown hyperactivity in bilateral amygdala, right putamen, anterior cingulate cortex, right insula, ventromedial prefrontal cortex, and right caudate extending to temporal and occipital cortices during symptom provocation (Thorsen, Hagland, et al. 2018; Yu et al. 2022).

Few studies have compared the neural activation patterns during symptom provocation between OCD patients with different symptom profiles. Patients with predominant contamination symptoms, when presented with standardized OCD-related triggers, had altered activity of amygdala extending to frontal and subcortical structures, and patients with doubts/checking symptoms had increased hippocampal activation (Thorsen, Kvale, et al. 2018; Phillips et al. 2000; Mataix-Cols et al. 2004). The severity of taboo thought-related symptoms is found to correlate with hyperactivity in limbic/paralimbic structures, including amygdala, anterior insula, anterior cingulate cortex, and lateral prefrontal cortex in a moral dilemma situation, which could be attributed to the heightened moral sensitivity experienced by these patients (Harrison et al. 2012). It can be thus surmised that taboo thought-related OCD may be associated with a hyperactive limbic system.

A majority of the symptom induction-based fMRI studies employed standardized triggers, which could have affected the efficacy of provocation, as the content of symptoms greatly varies among individual patients. Individualized symptom provocation pictures are noted to be associated with stronger activation of OCD-relevant regions, with some symptom-specific activation (Baioui et al. 2013; Viol et al. 2019). Further, inclusion of OCD patients with mixed symptom profiles may have confounded the results, leading to conflicting findings. Studying patients presenting with distinctive symptom profiles using individualized symptom provocation task may throw more light on the distinctive neural correlates of symptom profiles of OCD.

The current study attempts to compare the neural correlates of emotional provocation in OCD patients with predominantly contamination and taboo thought symptoms. These symptom profiles have been chosen as they have been consistently found to be distinctive in most factor or cluster analysis studies with preliminary evidence for differential neuroimaging correlates (Batistuzzo et al. 2021; Thorsen, Kvale, et al. 2018; Yu et al. 2022). We adopted an exploratory approach to compare the neural activation and connectivity patterns of frontolimbic circuit encompassing dorsomedial prefrontal cortex, ventromedial prefrontal cortex, anterior cingulate cortex, anterior insula, amygdala, and hippocampus between OCD patients with these two distinct symptom profiles and healthy individuals while visualizing personalized disorder-specific triggers.

## 2. Materials and Methods

### 2.1 Participants

A total of 54 subjects, including 37 OCD patients and 17 healthy controls, participated in the study. Patients were recruited from the specialty OCD clinic of the National Institute of Mental Health and Neuro Sciences (NIMHANS), Bengaluru, India. All recruited patients were symptomatic and had a diagnosis of primary OCD without other comorbid major psychiatric disorders. Healthy controls (HC) were recruited from the community by word of mouth and comprised of students/staff of the institute or healthy caregivers of patients. All participants were right-handed, had at least twelve years of education with the ability to read/write English, and did not have any medical or neurological comorbidities or contraindications to undergo magnetic resonance imaging. The Institutional Ethics Committee approved the study, and written informed consent was obtained from all participants. A detailed description of research design and participant details are given in the Supplement.

### 2.2 Clinical and diagnostic assessments

The diagnosis and comorbidities were confirmed using Mini International Neuro-Psychiatric Interview (MINI 7.0.2) (Sheehan et al. 1998). For patients, the severity and profiling of OCD were evaluated by Yale-Brown Obsessive-Compulsive Scale (Y-BOCS) and Yale-Brown Obsessive-Compulsive Scale-Symptom Checklist (Y-BOCS-SC) (Goodman, Price, Rasmussen, Mazure, Fleischmann, et al. 1989; Goodman, Price, Rasmussen, Mazure, Delgado, et al. 1989). OCD patients were categorized into either contamination obsessions/washing compulsions (OCD-C) (n=19) or taboo thoughts, i.e., sexual or religious obsessions and related compulsions (OCD-T) (n=18) groups based on the predominant symptom dimension. We defined predominant symptom dimension as the clinical profile that is reported as principal current and lifetime symptoms and are present from the onset of illness reported as most time-consuming, and impairing. To verify the presence of other principal symptom clusters, the dimensional severity was evaluated by Dimensional Yale-Brown Obsessive-Compulsive Severity Scale (D-YBOCS) (Rosario-Campos et al. 2006). Subjects with concomitant principal symptoms in any other dimension were excluded. The presence and severity of anxiety and depressive symptoms were measured using Hamilton Anxiety Rating Scale (HAM-A) and Hamilton Depression Rating Scale (HAM-D), respectively (M. Hamilton 1959; Max Hamilton 1967).

### 2.3 Functional Magnetic Resonance Imaging (fMRI) Paradigm

#### 2.3.1 Stimuli selection and validation

All participants were exposed to a set of personalized OCD and neutral stimuli during scan employing Individualized Symptom Provocation (ISP) task. A detailed symptom hierarchy worksheet was created for individual OCD subjects, enumerating situations or triggers for the principal obsessive-compulsive symptoms. Pictures simulating the triggering situations were collected either from the subjects’ home environment or other open sources as suggested by the participant. These disorder-specific stimuli were rated by patients on a scale of subjective units of distress (SUD) ranging from 0-10. The personalized disorder-specific pictures collected for OCD subjects were pooled and used for HC whilst preserving confidentiality of patients. HC were evaluated for the associated distress for disorder-specific stimuli based on SUD. To ensure effective experimental provocation for OCD patients, stimuli with SUD between 5 and 9 were included in the final ISP task.

A set of neutral stimuli along with disorder-specific stimuli were piloted on an independent sample of OCD patients, and all the triggers were rated on a scale of 0-10 SUD. Contrasting neutral triggers were chosen from this Neutral set for all participants undergoing imaging after evaluating for SUD. The SUD for each stimuli included in the task was re-examined after scan for all participants to confirm persistent provocation. Additional information on the validation of stimuli set and description of the stimuli are given in the supplement.

#### 2.3.2 Individualized symptom provocation (ISP) task

Stimuli from both conditions were presented as blocks in a randomized sequence, and each block had three stimuli presented for 10 seconds with an inter-block interval of 2200 msec. Each block was repeated thrice in the task (see Figure 1c). Prior to the scan, subjects were instructed to imagine themselves in the situation while visualizing the images, appraise their thoughts/emotions, and not to actively divert their attention or perform mental compulsions. Participants confirmed the distress or anxiety of SUD >5 for each stimulus through a button press.

**Figure 1.**
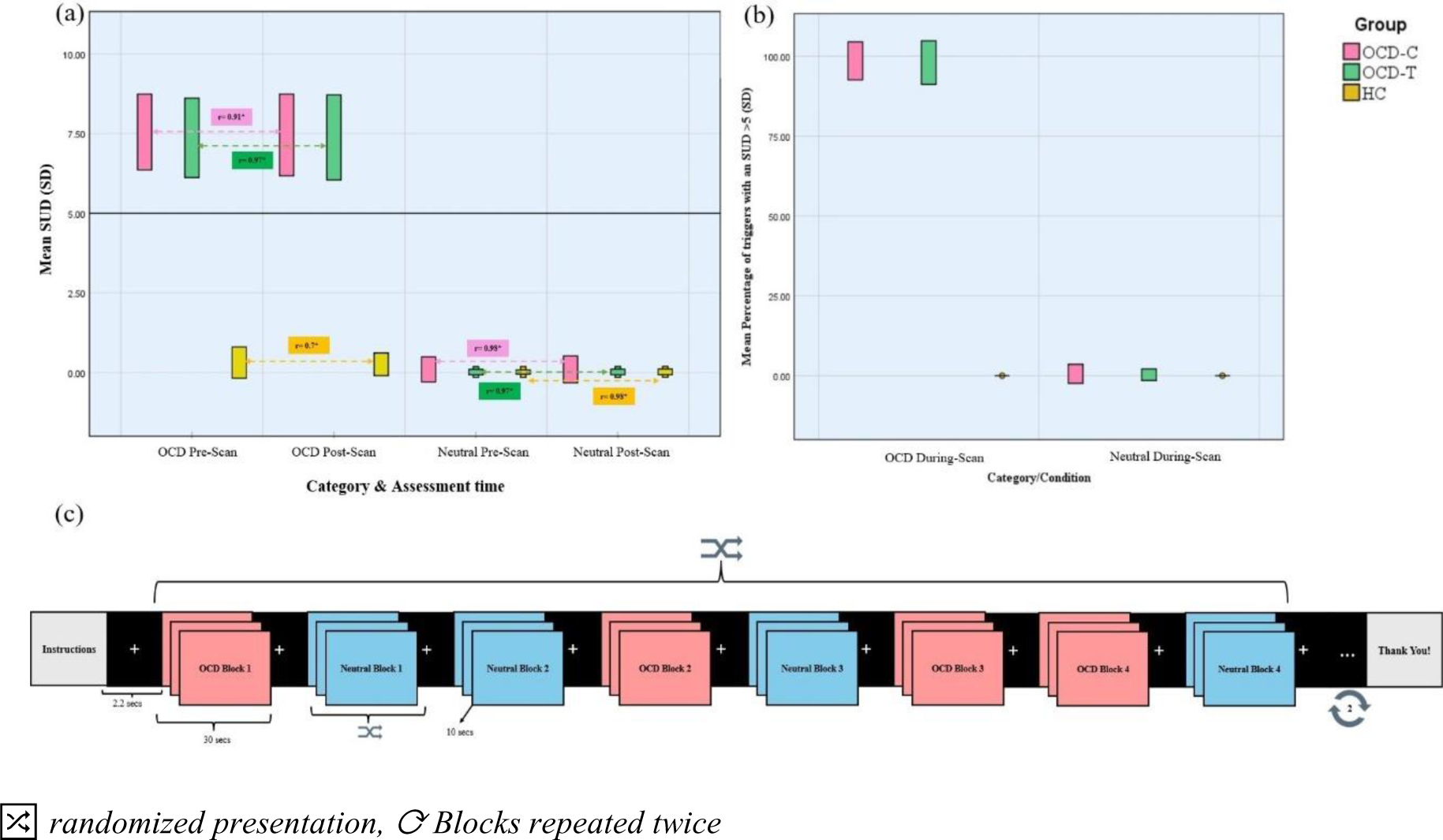
Individualized Symptom Provocation (ISP) Task (a) Distress ratings for the stimulus conditions before and after scan with correlation scores (b) Percentage of triggers from stimuli conditions reported to have a distress rating of >5 during scan and (c) fMRI paradigm design.

### 2.4 fMRI data acquisition

Magnetic resonance imaging data were acquired using a 3.0 Tesla scanner (Philips, Ingenia). High-resolution T1-weighted single-shot 3D turbo field echo (TFE) images were obtained with a repetition time (TR) of 2500 ms, echo time (TE) of 2.9 ms, 9° flip angle, field of vision (FOV) of 256×256×192 mm, slice thickness of 1 mm without inter-slice gap, 192 slices, matrix of 256×256 and voxel size of 1×1×1 mm. The blood oxygen level-dependent (BOLD) sensitive echo-planar sequences were acquired while subjects were performing the task with TR of 2200 ms, TE of 28 ms, 80° flip angle, FOV of 211×211mm, 3mm slice thickness, 44 slices, matrix of 64×62×64, and voxel size of 3.0 mm. The sequence lasted for 10 minutes, and 300 dynamic scans were obtained.

### 2.5 Clinical and behavioural data analyses

Sociodemographic and clinical variables across three groups were compared using Analysis of Variance (ANOVA) and post-hoc analyses are reported. The group-level comparisons of the mean distress (SUD) scores associated with OCD and neutral conditions before and after scans were performed by ANOVA with post-hoc analyses. The percentage of times distress/anxiety of SUD >5 was indicated for each condition was calculated and compared between groups using ANOVA. Group-wise Pearson correlation coefficient (r) was calculated to measure linear association between mean SUD scores for each condition before and after scan.

### 2.6 Imaging analyses

#### 2.6.1 fMRI data preprocessing and denoising

The fMRI was preprocessed using ENIGMA HALFpipe and fMRIPrep (Waller et al. 2022; Esteban et al. 2019). Preprocessing involved head motion estimation and correction, susceptibility distortion correction employing phase difference field maps, and outlier detection followed by coregistration and spatial normalization. Realignment of the functional data was performed where all dynamic scans were coregistered to the first scan of the session as a reference image employing a six-parameter rigid body transformation, and scans were resampled using b-spline interpolation to correct for motion and magnetic susceptibility interactions. Potential outliers were identified with framewise displacement above 0.5mm or global BOLD signal changes above five standard deviations. Functional and anatomical data were normalized into standard MNI space, segmented into grey matter, white matter, and CSF tissue classes, and resampled to 2 mm isotropic voxels following a direct normalization procedure. Final preprocessed functional images were obtained after the application of smoothing with a 6mm full-width half maximum (FWHM) kernel and grand mean scaling of 1000.

Noise reduction was performed by implementing anatomical component-based noise correction (aCompCor) procedure by regression of potential confounding effects from cerebral white matter, cerebrospinal fluid, head-motion parameters, outlier scans, and session and task effects with each factorial run. From the number of noise terms included in this denoising strategy, the effective degrees of freedom of the BOLD signal after denoising were estimated to range from 61.9 to 116 (average 100.5) across all subjects. Quality assessments to check for normalization, registration, and motion artifacts were performed on the final preprocessed functional images prior to first-level and group-level analyses.

#### 2.6.2 Region of interest (ROI) analyses

Anatomical masks were created on the MNI152NLin2009cAsym template with Harvard-Oxford Atlas parcellation (templateflow, Version 0.1.9, May 2019) (Ciric et al. 2022) based on the voxel coordinates for bilateral brain regions of ventromedial prefrontal cortex, dorsomedial prefrontal cortex, amygdala, hippocampus, anterior cingulate cortex, and anterior insula. A 5mm sphere was created around the reference voxel, and these selected voxels within the sphere were binarized (Figure S4).

Confirmatory analyses were conducted on FSL fMRI expert analysis tool (FEAT) (Woolrich et al. 2001). Thresholded statistical images were generated with contrast OCD>Neutral for each participant using timing data as an explanatory variable. Average beta aggregates from these anatomical seeds were interrogated from the statistical images of individual participants. Group-level comparisons of the mean activity across frontolimbic structures were performed employing an Analysis of covariance (ANCOVA) model after controlling for the age and sex distribution, homogeneity of the groups was evaluated with Levene’s test for equality of variances, and post hoc analyses were conducted with Bonferroni correction.

#### 2.6.3 Task-modulated functional connectivity analyses

The task-dependent effective connectivity between the hypothesized region of interest to every voxel was measured employing the generalized psychophysiological interference (gPPI) model on CONN functional connectivity toolbox version 22a (*CONN functional connectivity toolbox: RRID SCR_009550, release 22.* Hilbert Press. doi:10.56441/hilbertpress.2246.5840) (O’Reilly et al. 2012; Whitfield-Gabrieli and Nieto-Castanon 2012). Physiological regressor employed was the averaged BOLD time series extracted from the above-mentioned 11 Harvard-Oxford atlas ROIs included as seed regions and psychological regressor was the task effect calculated for the contrast of OCD>Neutral convolved with the hemodynamic response function. Functional connectivity changes across conditions were characterized by the multivariate regression coefficient of the psychophysiological interaction terms in each model.

Group-level comparisons were done using a General Linear Model (GLM) with each voxel as a dependent variable and groups of subjects as the independent variable after controlling for the effect of age and sex. Voxel-level hypotheses were evaluated using multivariate parametric statistics with random effects across subjects and sample covariance estimation across multiple measurements. Inferences were performed at the level of individual clusters (groups of contiguous voxels). Cluster-level inferences were based on parametric statistics from Gaussian Random Field theory. Results were thresholded using a combination of a cluster-forming p <0.001 voxel-level threshold and a false discovery rate corrected (p-FDR <0.05) cluster-size threshold (Nieto-Castanon 2020). Additional sensitivity analyses are elaborated in the Supplement.

## 3. RESULTS

### 3.1 Demographic and clinical measures

The subjects across three groups were comparable in all socio-demographic domains except for sex (Table 1). The OCD-T group reported more anxiety and depression symptoms than the other two groups on clinical assessments (Figure S3).

**Table 1.** Sociodemographic and clinical characteristics of the sample.

| **Sociodemographic and clinical variable** | **OCD-C**<br>**(n=19)** | **OCD-T**<br>**(n=18)** | **Healthy**<br>**controls (n=17)** | **p-value**<br>**(corrected)** |
| --- | --- | --- | --- | --- |
| Age at recruitment mean in years (SD) | 30.32 (8.18) | 27.44 (6.9) | 30.59 (7.79) | 0.4 |
| Sex - males, n(%) | 6 (31.6) | 15 (83.3) | 10 (58.8) | **0.006** |
| Years of education mean (SD) | 13.89 (3.7) | 15.4 (3.94) | 16.76 (3.63) | 0.08 |
| HAM-A scores estimated mean* (SE) | 5.25 (0.67) | 9.38 (0.7) | 1.25 (0.6) | **<0.001** |
| HAM-D scores estimated mean* (SE) | 4.55 (0.63) | 7.47 (0.65) | 0.54 (0.62) | **<0.001** |
| Family history of OCD n (%) | 3 (15.8) | 1 (5.6) | 0 | 0.183 |
| Total duration of illness in years, mean (SD) | 9.66 (6.83) | 10.75 (6.93) | - | 0.63 |
| Age at onset mean (SD) | 20.7 (5.06) | 16.6 (7.85) | - | 0.06 |
| Total Y-BOCS mean (SD) | 29.26 (5.85) | 27.56 (4.93) | - | 0.35 |
| Treatment status (currently on SRI)n (%) | 17 (89.5) | 15 (83.3) | - | 0.79 |
| SRI resistance<sup>+</sup> n (%) | 4 (21.1) | 3 (16.7) | - | 1 |

HAM-A- Hamilton Anxiety Rating Scale, HAM-D- Hamilton Depression Rating Scale, YBOCS- Yale-Brown Obsessive-Compulsive Scale, SRI- Serotonin Reuptake Inhibitor.
\*covariate (age, sex distribution) adjusted mean + failure to respond to $\geq 2$ adequate trials of SRIs

### 3.2 Behavioral measures

The mean distress reported for OCD stimuli pre- and post-scan were comparable between patient sub-groups. Healthy controls experienced minimal provocation with disorder-specific stimuli before and after scan (Figure 1(a) and Table S3). Out of 36 OCD stimuli visualized during the scan, SUD >5 was reported by OCD-C group on an average of 98.54% (SD: 2.98) and OCD-T group for 97.9% (SD: 3.41) (Figure 1(c)). A significant linear correlation was noted between the SUD scores of both the conditions, pre- and post-scans (Figure 1(a) and Table S4).

### 3.3 Task-based activation

The between-group difference for the task contrast of OCD>Neutral is illustrated in Table S5. OCD subjects with contamination symptoms were noted to have significantly reduced activity in left amygdala during symptom provocation than those with taboo thought symptoms (adjusted mean difference= 15.68, *p*(Bonferroni corrected)= 0.01), and healthy controls (adjusted mean difference= 13.48, *p*(Bonferroni corrected)= 0.03). Contamination group was also noted to have reduced right hippocampus mean activity as compared to healthy individuals (adjusted mean difference= 17.05, *p* (Bonferroni corrected)= 0.04) while visualizing disorder-specific triggers over neutral triggers (Figure 2).

**Figure 2.**
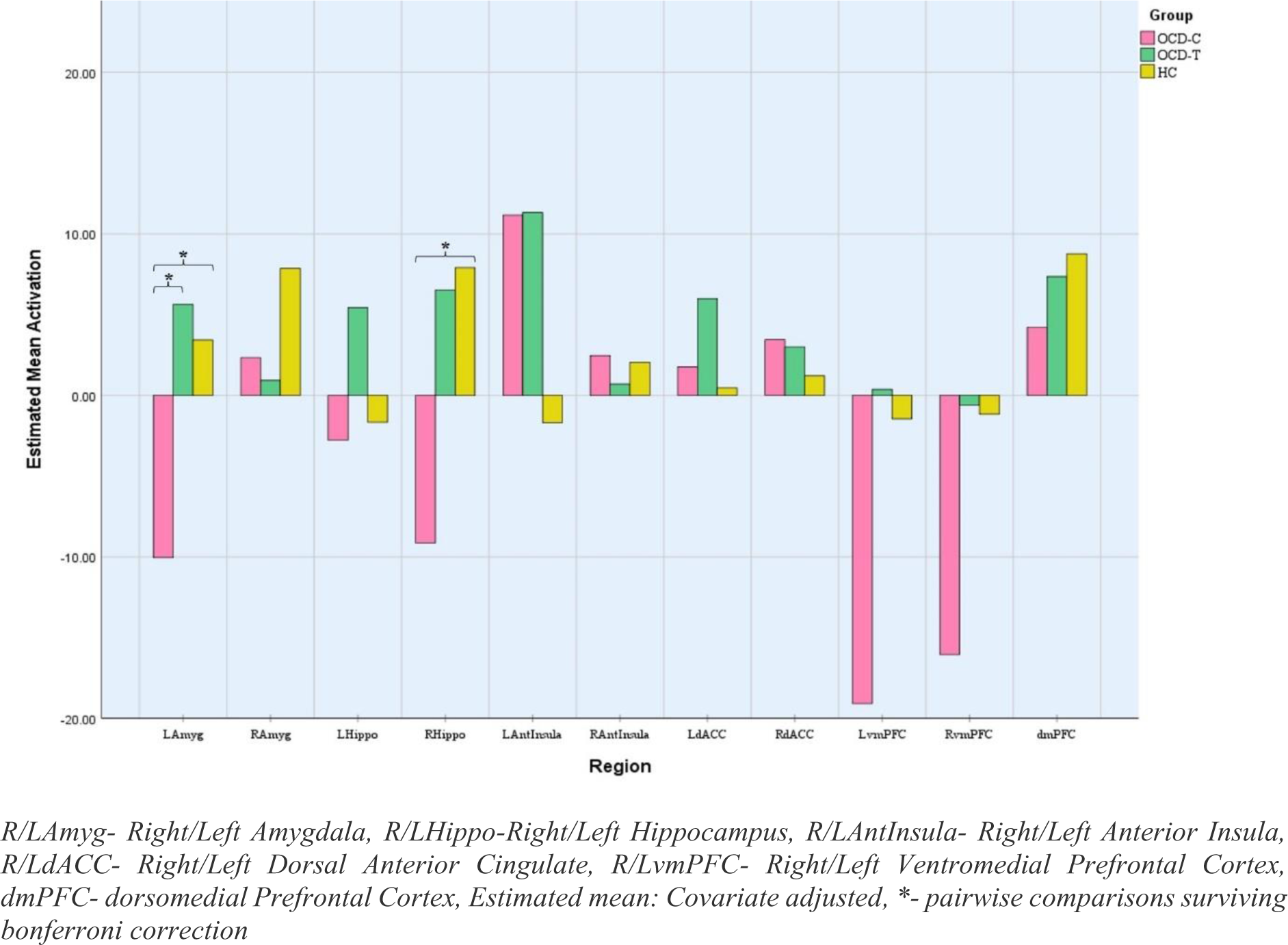
Task-based estimated mean activation during symptom provocation state (OCD>Neutral)

### 3.4 Task-modulated functional connectivity

The significant group-level comparisons of the whole-brain task-based connectivity changes are depicted in Table 2. The OCD-T subjects had greater connectivity between right insula and left cerebellum than OCD-C group (T= 6.64, p-FDR= <0.001) and increased connectivity between medial prefrontal cortex and superior division of right lateral occipital cortex than healthy controls (T= 5.08, p-FDR= <0.001) while visualizing disorder-specific over neutral stimuli. Healthy subjects had greater connectivity of the right hippocampus with bilateral supplementary motor area and anterior cingulate gyrus (T= 5.11, p-FDR= 0.04), and bilateral insular cortex with left middle temporal cortex, left cerebellum and bilateral intracalcarine cortex while appraising provocative triggers over disorder-specific stimuli than subjects with contamination symptoms (right insula: T= 5.47, and p-FDR= 0.02, T= 4.55, p-FDR= 0.03; left insula: T= 6.27, p-FDR= 0.03) (Figure 3).

**Figure 3.**
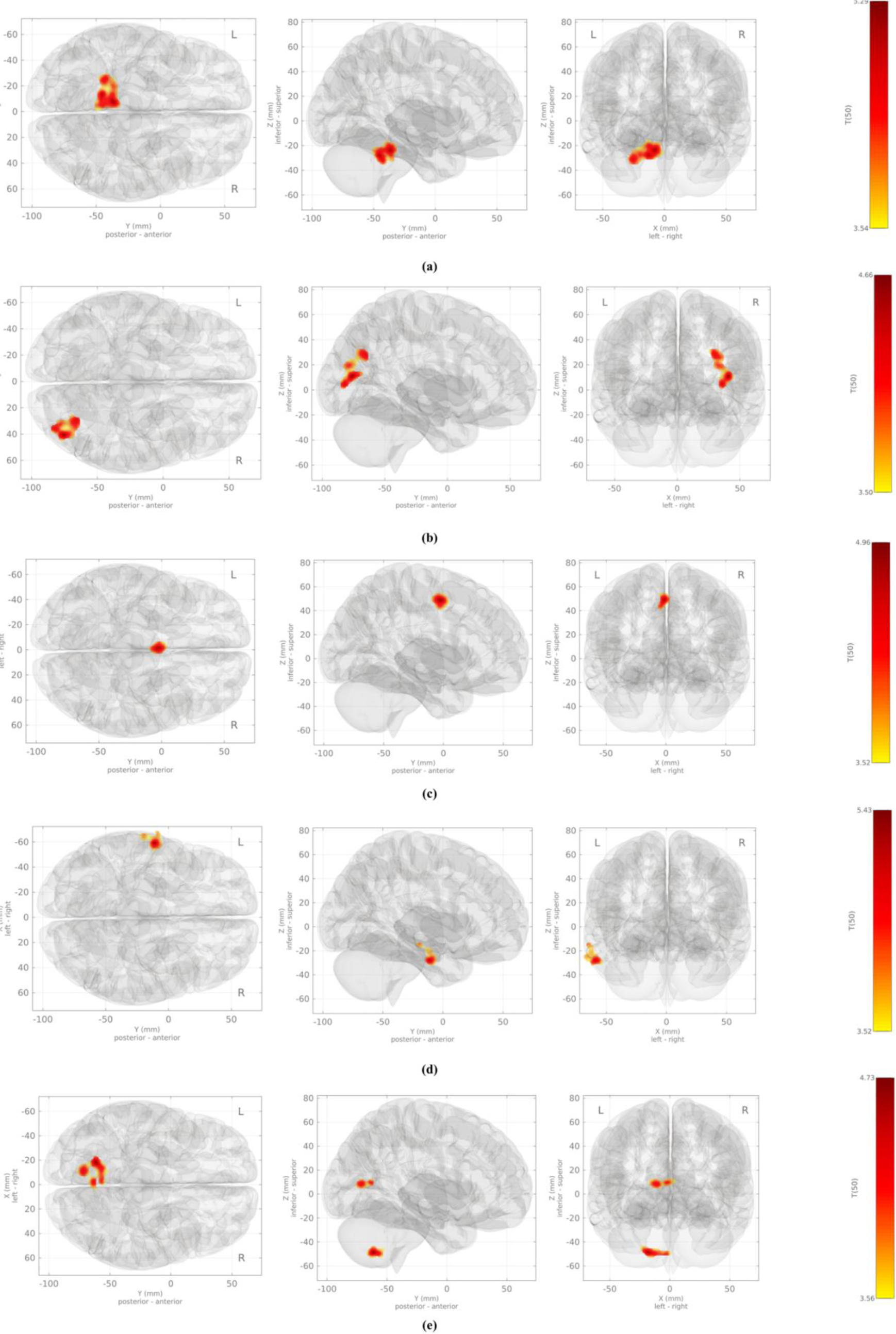
Network maps depicting the task-based functional connectivity of frontolimbic structures during emotionally provoked condition (OCD>Neutral) (a) Right insula: OCD- T>OCD-C; (B)Medial frontal cortex: OCD-T>HC; (C) Right hippocampus: HC>OCD-C; (D) Left insula: HC>OCD-C; (E) Right insula: HC>OCD-C.

**Table 2.** Task-modulated (OCD>Neutral) connectivity of frontolimbic circuit.

| Contrast | Seed | Target | Cluster size | T-value | p-FDR corrected |
| --- | --- | --- | --- | --- | --- |
| OCD-T>OCD-C | Right Insular cortex | Left cerebellum and vermis | 396 | 6.64 | <0.001 |
| OCD-T>HC | Medial frontal cortex | Right lateral occipital cortex, superior and inferior division | 232 | 5.08 | <0.001 |
| HC>OCD-C | Right Hippocampus | Bilateral Supplementary Motor Cortex, Anterior cingulate gyrus | 100 | 5.11 | 0.04 |
|  | Left Insular cortex | Left middle and inferior temporal gyrus, anterior and posterior division | 105 | 6.27 | 0.03 |
|  | Right Insular cortex | Left Posterior Cerebellum | 115 | 5.47 | 0.02 |
|  |  | Bilateral Intracalcarine cortex | 93 | 4.55 | 0.03 |

*p*-FDR corrected: <0.05

## 4. DISCUSSION

We investigated the effect of emotional processing of personalized disorder-specific triggers on the functional activation and connectivity of the frontolimbic circuit among OCD subjects with two well-delineated symptom profiles, i.e., contamination and taboo thought-related symptoms. Individualized stimuli are considered superior in eliciting stronger and discrete emotional reactions associated with specific symptom expression. Provocation with highly personalized stimuli, as compared to standardized disorder-specific and neutral triggers, causes increased activation of amygdala, hippocampus, cingulate cortex, and medial frontal cortex among OCD patients with mixed presentation correlating to self-referential processing and memory retrieval (Viol et al. 2019). Our findings suggest that the appraisal of personalized OCD compared to neutral stimuli caused reduced activation and connectivity of various frontolimbic structures in OCD patients with contamination concerns than those with taboo thoughts or healthy individuals.

Left amygdala has an established role in the rapid processing of image- or language-related affective information, and activation of amygdala is hypothesized to facilitate representation and retrieval of emotional memory by hippocampus (Daum, Markowitsch, and Vandekerckhove 2009; Phelps and Ledoux 2000; Qasim et al. 2023). Reduced activation indicates an inadequate emotional evaluation and registration of the provocative stimuli by the subjects with contamination dimension. Additionally, aberrant connectivity between hippocampus and SMA can indicate inefficient utilization of retrieved emotional memory, leading to abnormal motor planning with abnormalities in execution of complex behaviors based on the inputs from the environment (van den Heuvel et al. 2016; Shephard et al. 2021; 2022).

Altered hippocampal-anterior cingulate gyrus connectivity can be speculated to be associated with abnormal error monitoring, cognitive flexibility, and emotional regulation, frequently noted in OCD patients (Fitzgerald et al. 2005; Gonçalves et al. 2016; McGovern and Sheth 2017). Recent work has suggested that the fronto-hippocampal interactions, including that from the SMA and dACC, are involved in retrieval stopping, i.e., inhibition of unpleasant emotional memories (Anderson and Floresco 2022). Overall, an altered interaction between these components of frontolimbic and sensorimotor circuits could potentially mediate the influence of abnormal emotional processing, leading to disrupted cognitive control of the prepotent automated motor responses and the emergence of compulsive behaviors (van den Heuvel et al. 2016).

The middle and inferior temporal gyrus (MTG/ITG) facilitate semantic memory processing, visual perception, and integration of multimodal sensory inputs (Mesulam 1998; Herath, Kinomura, and Roland 2001; Wei et al. 2012). Reduced activity of the left MTG among OCD subjects was reported to be associated with poor insight, reflecting probable deficits in memory encoding and retrieval (Fan et al. 2017), and patients with contamination symptom profiles were reported to have significantly impaired visuospatial recognition memory and working memory than other symptom dimensions (H. Kashyap et al. 2017; Dittrich et al. 2011). Additionally, cerebellum has a pivotal role in emotional processing learning (Baumann and Mattingley 2022; Adamaszek et al. 2017). The altered cerebellar activity is hypothesized to be a compensatory phenomenon secondary to cortico-striatal dysconnectivity in OCD (R. Kashyap et al. 2021; Zhang et al. 2019).

OCD patients experience behavioral inflexibility, impaired cognitive control, and abnormal error monitoring due to aberrant functioning of dorsal and ventral MFC (Gillihan et al. 2011; Simon, Rudebeck, and Rich 2021; Yücel et al. 2007). The midline structures, including MFC, right lateral occipital gyrus and other visual association areas, are involved in self-referential processing, and hyperactivation of cortical midline structures is associated with self-referential tasks in OCD patients (Viol et al. 2019; Northoff et al. 2006; Hu et al. 2016). The increased activity of MFC and visual association areas reflect heightened visual processing of stimuli with increased involvement of self-referential thinking during emotional provocation, presumably due to associated anxiety.

In summary, these findings from our sample of OCD patients with one predominant symptom dimension devoid of major psychiatric or medical comorbidities suggest that contamination dimension is associated with deficiencies in emotional memory formation leading emotional regulation abnormalities and taboo thoughts are accompanied by enhanced emotional evaluation and awareness.

### Limitations

These findings must be interpreted cautiously, as sample size was limited, which could have reduced the statistical power to identify other important findings. Secondly, as most of the OCD subjects were on psychotropics, we could not control for the potential effects of medications on neural activation patterns. Further, we employed disorder-specific stimuli from the personal environment of OCD subjects to compare neural activation in healthy individuals. Thus, the effect of novelty with these stimuli in healthy controls could not be controlled considering the self-referential significance of the triggers among OCD subjects. However, the distinctive reduced connectivity observed in the contamination group alone and not in the taboo thoughts group in contrast to healthy individuals hints that these findings might be related to symptom provocation rather than familiarity.

In spite of our attempt to maintain the homogeneity of the included sample, the possibility of heterogeneity within a given symptom dimension can not be disregarded. Contamination obsessions and washing/cleaning compulsions can manifest secondary to fear, avoidance, perfectionism, disgust sensitivity, or sensory phenomena. Thus, deep phenotyping of symptom dimensions on larger samples might help to obtain insight into this complex disorder. Understanding the distinctive correlates of symptom dimensions can improve our insight into the etiopathogenesis of this heterogeneous disorder and pave the way for developing personalized/targeted neuromodulatory interventions.

## Data Availability

The imaging and clinical data in the present study will be available upon reasonable request to the corresponding author.

## Acknowledgements

NST acknowledges the salary support received from the Department of Biotechnology, Ministry of Science and Technology, Government of India funded project “Accelerator Program for Discovery in Brain disorders using Stem cells” (ADBS) (BT/PR17316/MED/31/326/2015). SSA has received a research grant from DBT/Wellcome Trust India Alliance (IA/CPHI/18/1/503931).

