## Supplement for "Differential connectivity of frontolimbic circuit induced by individualized disorder-specific stimuli in distinct symptom profiles of obsessive-compulsive disorder"

**Pilot study for validating fMRI individualized symptom provocation (ISP) task:**

A pilot study was done on an independent sample of OCD patients with either principal contamination/washing or aggressive/doubts/checking or taboo-thought symptoms (n= 32 [female= 21], mean age of 39.2 years, mean YBOCS: 24.2) and healthy individuals (n=19 [female=13], mean age of 35.72 years) to standardize the method of selection of individualized disorder-specific triggers for the final fMRI task (Figure S1) and validate contrasting set of neutral stimuli set (*Tendler A et al., 2019*). For OCD stimuli, a hierarchy of situations or triggers provoking oc-symptoms was created for individual OCD subject, along with details of the obsessions, subjective units of distress (SUD) associated on a scale of 0 to 10 and subsequent urge to perform a compulsive behaviour. Images simulating the situation or trigger was collected with the support of OCD subjects and SUD was re-evaluated for the stimuli employing cognitive appraisal strategy. Pictures without any confidentiality concerns were pooled and healthy individuals rated these stimuli on a similar scale of SUD. We ensured that subjects will not be exposed to explicit sexually-provoking or grossly disgust-inducing stimuli employing independent clinicians to rate these stimuli.

Following this, a similar strategy was employed to generate a set of neutral stimuli that did not induce distress among subjects. Any picture in the neutral category provoking distress of more than 5 SUD was excluded from the final set. A set of twenty-five pictures were selected for the neutral condition. The selected visual stimuli from both conditions were resized and normalized for contrast and saturation before generating the fMRI task. The OCD-C symptom triggers can be categorized as themes of contamination by feces, dirt, blood stains, garbage, unclean dishes, street foods/vendors, sanitation workers, food stuck on gums/teeth/nails, mucosal discharge, seminal discharges, insects including cockroaches or houseflies, soiling of clothes with fecal or menstrual blood stains, stains on toilet seats or washbasins. Broadly, theme of taboo-thought triggers included couples (homosexual/heterosexual) hugging/kissing, pictures of children playing/sobbing, women dressed in slightly revealing indian wear (saree), paintings of hindu gods/goddesses, ancient hindu idols, sculptures from khajuraho, pictures of mosques or churches or temples in the neighborhood of individual OCD subjects’, pictures with objects of religious importance. Providing the details of personalized triggers from OCD-T category is deliberately avoided to protect confidentiality of participants. Description of the neutral stimulus set used in the fMRI task are provided Table S1.

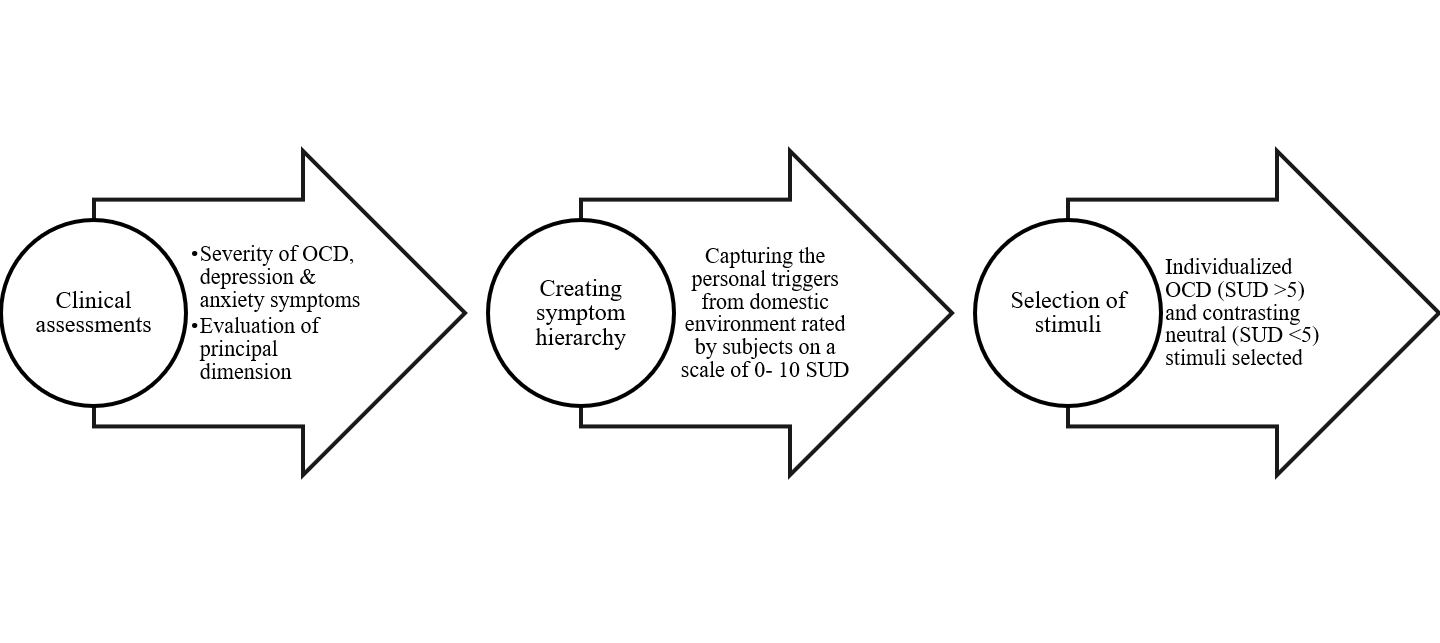
Figure S1 Method of personalized stimuli selection for individualized symptom provocation paradigm

Table S1 Description of neutral stimuli set with average SUD rating by the pilot sample

| Description of Neutral stimuli | Mean SUD*  OCD group (n= 32) | Mean SUD*  HC group (n=19) |
| --- | --- | --- |
| Lake during sunset | 0 | 0 |
| Butterfly | 0 | 0 |
| Lake with reflection of mountains | 1.2 | 0 |
| Sparrow | 0.8 | 0 |
| Sunflowers | 0 | 0 |
| Mother and baby giraffe | 0.7 | 0.06 |
| Cloudy skies during sunset | 1.6 | 0 |
| Cherry blossom | 0 | 0 |
| Assorted grain bags | 1.1 | 0 |
| Table lamp | 0.9 | 0 |
| Fruit shop | 0.6 | 0 |
| Person painting a flower (hind view) | 0 | 0 |
| Chairs in a garden with cloudy skies backdrop | 0.2 | 0 |
| Train in snowy mountains | 0.8 | 0 |
| Corn fields | 1.3 | 0.02 |
| Fish underwater | 0.9 | 0.07 |
| Helicopter | 1.9 | 0 |
| Flowers and butterfly | 0 | 0 |
| Field with cloudy skies | 0.2 | 0 |
| Boat in clear water | 0 | 0 |
| Chair in the corner of a room | 0.4 | 0 |
| Lake with reflection of clouds | 0 | 0 |
| Peanuts | 2.1 | 0 |
| Flowers in a garden | 0 | 0 |
| Spoon on a wooden table | 0 | 0 |

*Mean Subjective units of distress (SUD) on a scale of 0 to 10

Table S2 Selection Criteria

| Group | Specific Inclusion Criteria | Common Inclusion Criteria | Specific Exclusion Criteria | Common Exclusion Criteria |
| --- | --- | --- | --- | --- |
| **OCD- C (predominant contamination/washing)** | Diagnosis of OCD- MINI 7.0.2, predominant contamination obsessions/ Washing compulsions- YBOCS-SC and no other principal symptom dimensions, YBOCS severity score ≥16, CGI-S score ≥4 | 1. Age range: 18-45 years 2. Males or females 3. At least 10 years of formal education 4. Able to read and write English 5. Right-handed as assessed with Edinburgh Handedness Inventory | 1. Lifetime diagnosis of other Bipolar Affective Disorder, Schizophrenia, psychotic disorders as assessed by MINI.7.0.2. 2. Substance use disorders except nicotine dependence syndrome. 3. Current severe major depressive disorder | 1. Co-existing major neurological illnesses-CVA, epilepsy, demyelinating diseases, neuro-infections, head injuries or history of neurosurgeries, delayed development milestones. 2. Pregnancy, post-partum/lactating women 3. Contraindications to MRI (Cardiac pacemakers, aneurysm clips, cochlear implants, history of metal fragments in eyes, claustrophobia, orthodontic braces and others). |
| **OCD-TT (Predominant taboo thoughts)** | Diagnosis of OCD- MINI 7.0.2, predominant taboo thoughts- sexual/religious obsessions- YBOCS-SC and no other principal symptom dimensions, YBOCS severity score ≥16, CGI-S score ≥4 |  |  |  |
| **Healthy controls (HC)** | **-** |  | Any identifiable psychiatric disorder- MINI 7.0.2 |  |

YBOCS: Yale-Brown Obsessive-Compulsive Scale, CGI-S: Clinical Global Impression- Severity Scale; MINI 7.0.2: Mini International Neuro-Psychiatric Interview (MINI 7.0.2); CVA: Cerebrovascular accidents, MRI: Magnetic Resonance Imaging

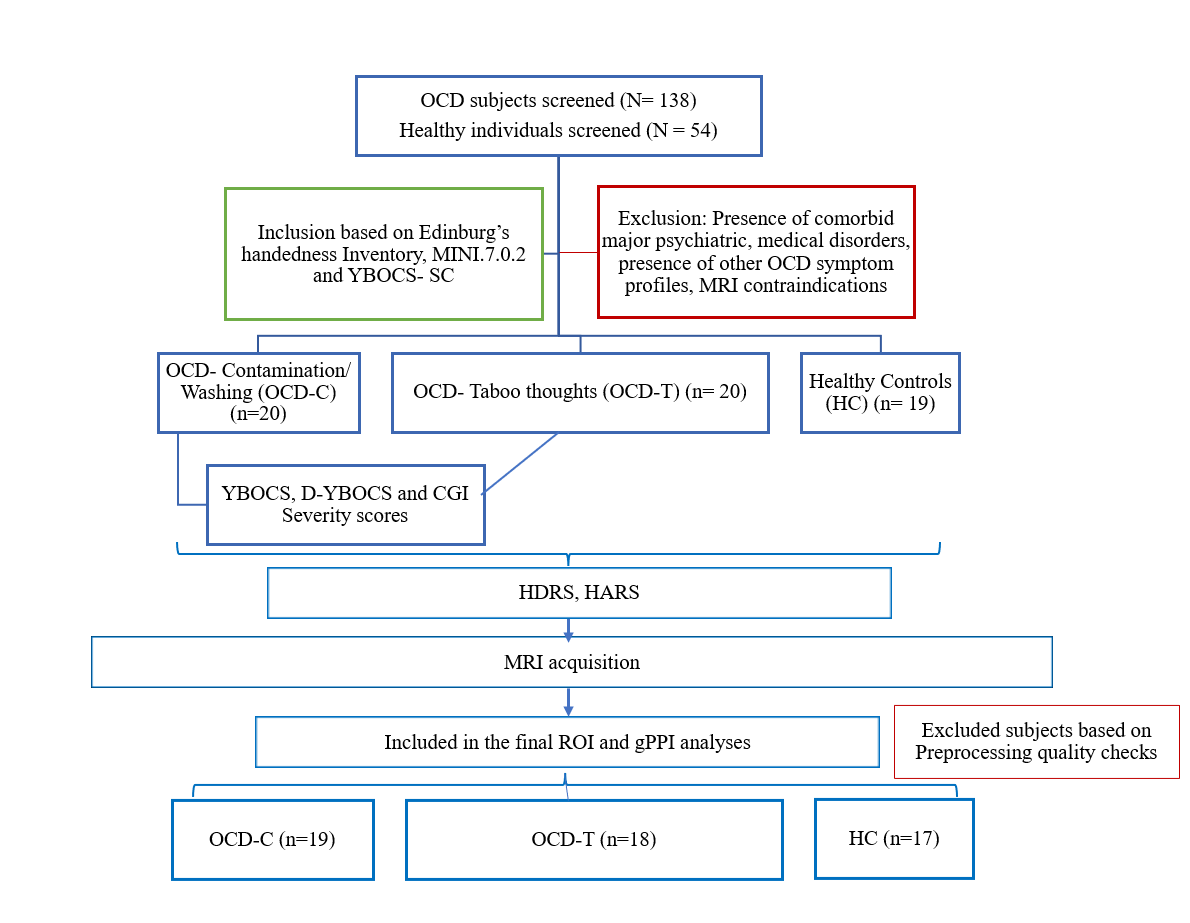

Figure S2 Flowchart of inclusuion of subjects for final analyses

MINI 7.0.2- Mini International Neuropsychiatric Interview version, YBOCS- Yale-Brown Obsessive-compulsive scale, Severity Scale, SC- Symptom Checklist, D-YBOCS- Dimensional Yale-Brown Obsessive Compulsive Scale, CGI-S -Clinical Global Impressions-Severity scale, ROI- Region of Interest, gPPI: generalized Psychophysiological interference

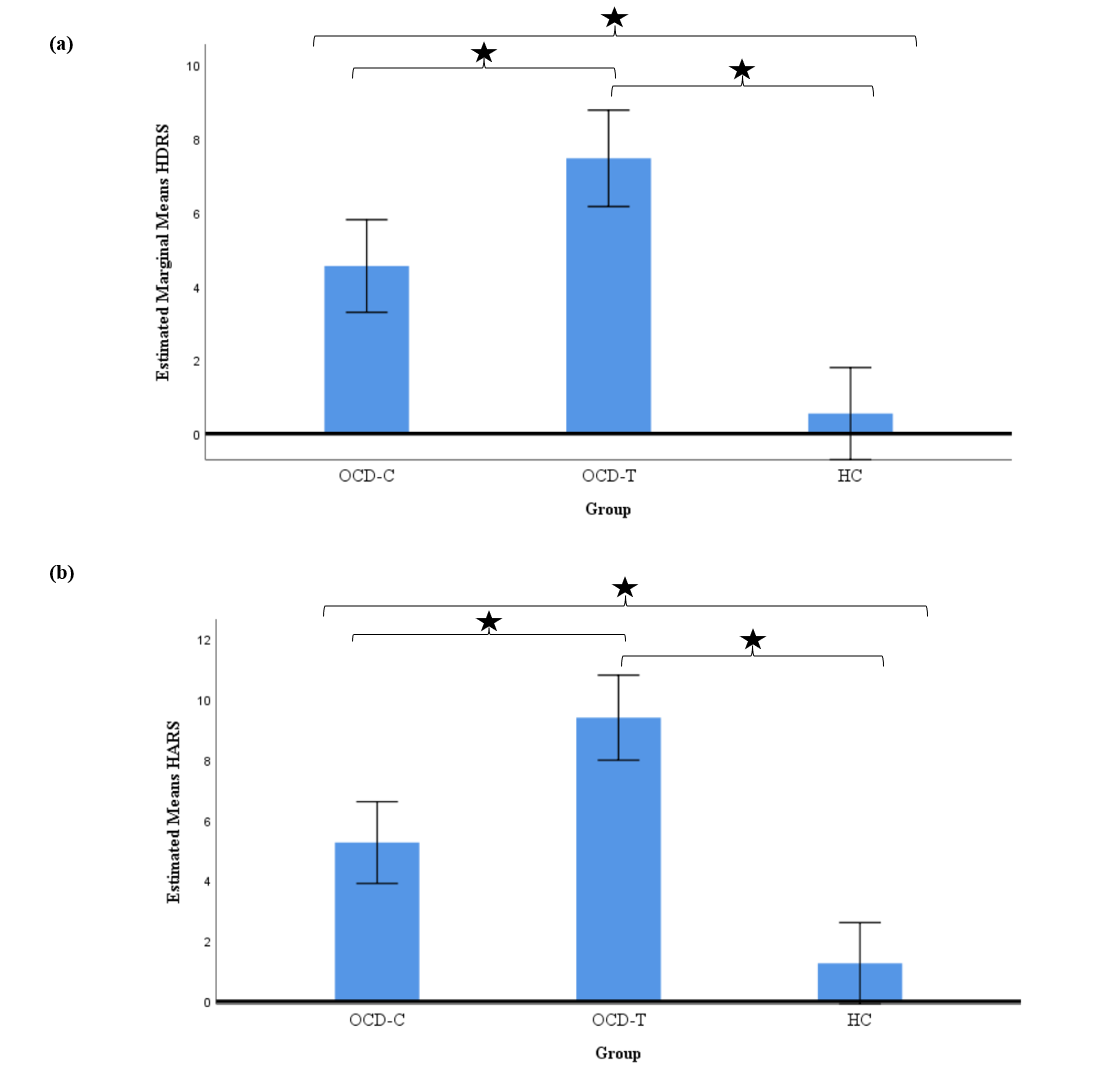

Figure S3 Three-group comparison of (a) Hamilton Depression Rating Scale scores (F (2,54)= 30.31, *p=* <0.00) and (b) Hamilton Anxiety Rating Scale scores (F (2,54)= 35.04, *p*= <0.00) after controlling for age and sex distribution.

Table S3 Behavioural measures

| **SUD: Mean(SD)** | **OCD-C (n=19)** | **OCD-T (n=18)** | **HC (n=17)** | ***p-value*** |
| --- | --- | --- | --- | --- |
| **PreScan-OCD** | 7.55 (0.6) | 7.37 (0.62) | 0.31 (0.24) | ***<0.001*** |
| **PostScan-OCD** | 7.46 (0.64) | 7.38 (0.67) | 0.26 (0.18) | ***<0.001*** |
| **PreScan-Neutral** | 0.1 (0.2) | 0.01 (0.04) | 0.01 (0.03) | 0.068 |
| **PostScan-Neutral** | 0.1 (0.22) | 0.02 (0.04) | 0.02 (0.04) | 0.103 |
| **Percentage of OCD stimuli with SUD >5 duringScan (N=36)** | 98.54 (2.98) | 97.9 (3.41) | 0 (0) | ***<0.001*** |
| **Percentage of Neutral stimuli with SUD >5 duringScan (N=36)** | 0.59 (1.49) | 0.31 (0.9) | 0 (0) | 0.241 |

SUD: Subjective Units of Distress

Table S4 Group-level correlation matrix between pre- and post-scan SUD for OCD and Neutral conditions

| **Group** | Category | PreScanOCD (r, p-value) | PostScanOCD (r, p-value) | Category | PreScanNeutral (r, p-value) | PostScanNeutral (r, p-value) |
| --- | --- | --- | --- | --- | --- | --- |
| **Con/Wash** | PreScanOCD | 1 | 0.91 **(<0.00)** | PreScanNeutral | 1 | 0.98 (<0.00) |
| **TT** | PreScanOCD | 1 | 0.97 **(<0.00)** | PreScanNeutral | 1 | 0.97 (<0.00) |
| **HC** | PreScanOCD | 1 | 0.7 **(0.002)** | PreScanNeutral | 1 | 0.98 (<0.00) |

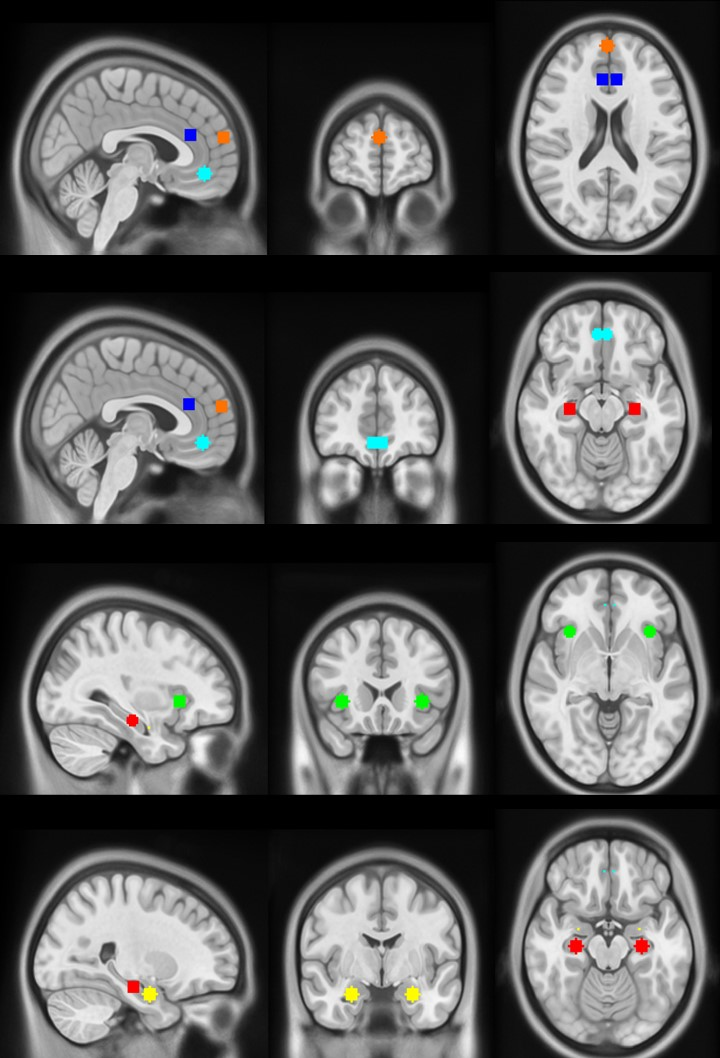

Figure S4 Seeds for the region of interest (ROI) analysis depicted on MNI152NLin2009cAsym T1-w template. Light blue: ventromedial prefrontal cortex, orange: dorsomedial prefrontal cortex, deep blue: anterior cingulate cortex, green: anterior insula, red: hippocampi, yellow: amygdala

Table S5 Mean activation of the frontolimbic circuit during symptom provocation (OCD>Neutral)

| **Region** | ***Estimated mean (sd)** | | | ****Pairwise comparisons** | | | **F-statistic** | ***p-value** |
| --- | --- | --- | --- | --- | --- | --- | --- | --- |
|  | **OCD-C** | **OCD-T** | **HC** | ****Significant group difference** | ****Mean difference** | ****p-value** |  |  |
| Left amygdala | -10.05 (3.57) | 5.63 (3.71) | 3.43 (3.55) | OCD-T>OCD-C | 15.68 | ***0.01*** | 5.09 | ***0.01*** |
|  |  |  |  | HC>OCD-C | 13.48 | ***0.03*** |  |  |
| Right amygdala | 2.34 (4.68) | 0.93 (4.87) | 7.87 (4.65) | - | - | - | 0.62 | 0.53 |
| Left hippocampus | -2.77 (4.60) | 5.43 (4.78) | -1.66 (4.58) | - | - | - | 0.80 | 0.45 |
| Right hippocampus | -9.14 (4.72) | 6.52 (4.91) | 7.91 (4.7) | HC>OCD-C | 17.05 | ***0.04*** | 3.77 | ***0.03*** |
| Left Anterior Insula | 11.16 (4.7) | 11.32 (4.85) | -1.68 (4.64) | - | - | - | 2.65 | 0.08 |
| Right Anterior Insula | 2.47 (4.83) | 0.71 (5.02) | 2.05 (4.8) | - | - | - | 0.03 | 0.97 |
| Left anterior cingulate cortex | 1.77 (4.73) | 5.99 (4.92) | 0.46 (4.7) | - | - | - | 0.34 | 0.71 |
| Right anterior cingulate cortex | 3.45 (4.4) | 3.01 (4.57) | 1.21 (4.37) | - | - | - | 0.08 | 0.92 |
| Left ventromedial prefrontal cortex | -19.08 (5.46) | 0.37 (5.68) | -1.45 (5.43) | - | - | - | 3.52 | **0.03** |
| Right ventromedial prefrontal cortex | -16.04 (5.1) | -0.6 (5.31) | -1.17 (5.07) | - | - | - | 2.7 | 0.07 |
| Dorsomedial prefrontal cortex | 4.22 (6.52) | 7.36 (6.77) | 8.77 (6.48) | - | - | - | 0.13 | 0.88 |

*covariate (sex distribution) adjusted ***p*-Bonferroni corrected <0.05

| **Contrast** | **Seed** | **Target** | **Cluster size** | **T-value** | **p-FDR corrected** |
| --- | --- | --- | --- | --- | --- |
| OCD-T>OCD-C | Right Insular cortex | Left cerebellum and brain stem | 212 | 5.91 | <0.001 |
| OCD-C>OCD-T | Left Hippocampus | Not labelled* | 131 | 5.75 | 0.013 |
| OCD-T>HC | Left Amygdala | Supracalcarine cortex, precuneous | 152 | 4.72 | 0.003 |
|  |  | Left frontal pole | 81 | 4.78 | 0.02 |
|  |  | Precuneous | 77 | 4.66 | 0.02 |
| HC>OCD-C | Right Amygdala | Left occipital cortex, left precuneous, left intracalcarine cortex | 156 | 5.21 | 0.006 |
|  |  | Right intracalcarine cortex, right occipital cortex, right supracalcarine cortex | 124 | 4.72 | 0.01 |
|  | Right Insular cortex | Left Posterior Cerebellum | 110 | 5.03 | 0.02 |
|  |  | Left Intracalcarine cortex, precuneous | 82 | 4.54 | 0.04 |

Table S6 Significant clusters of group-level comparisons of task-modulated (OCD>Neutral) connectivity of frontolimbic circuit after controlling for age, sex, anxiety and depression symptom severity

| **Contrast** | **Seed** | **Target** | **Cluster size** | **T-value** | **p-FDR corrected** |
| --- | --- | --- | --- | --- | --- |
| OCD-T>OCD-C | Right Insular cortex | Left cerebellum and brain stem | 184 | 6.76 | <0.001 |

Table S7 Task-modulated (OCD>Neutral) connectivity of frontolimbic circuit across OCD groups after controlling for age, sex, obsessive-compulsive, anxiety and depression symptom severity

Table S8 Correlation of dimensional contamination and taboo thought scores with task-modulated (OCD>Neutral) connectivity after controlling for age, sex, anxiety and depression symptoms

| **Contrast** | **Seed** | **Target** | **Cluster size** | **T-value** | **p-FDR corrected** |
| --- | --- | --- | --- | --- | --- |
| DYBOCS-TT> DYBOCS-Con | Right Insular cortex | Left cerebellum and brain stem | 328 | 6.6 | <0.001 |
|  | Medial frontal cortex | Left precentral gyrus | 114 | 5.75 | 0.04 |

DYBOCS-TT: dimensional scores for taboo thought symptoms, DYBOCS-Con: dimensional scores for Contamination/Washing symptoms
